# MOVER: Medical Informatics Operating Room Vitals and Events Repository

**DOI:** 10.1101/2023.03.03.23286777

**Authors:** Muntaha Samad, Joseph Rinehart, Mirana Angel, Yuzo Kanomata, Pierre Baldi, Maxime Cannesson

## Abstract

Artificial Intelligence (AI) holds great promise for transforming the healthcare industry. However, despite its potential, AI is yet to see widespread deployment in clinical settings in significant part due to the lack of publicly available clinical data and the lack of transparency in the published AI algorithms. There are few clinical data repositories publicly accessible to researchers to train and test AI algorithms, and even fewer that contain specialized data from the perioperative setting. To address this gap, we present and release the Medical Informatics Operating Room Vitals and Events Repository, which includes data from 58,799 unique patients and 83,468 surgeries collected from the UCI Medical Center over a period of seven years. MOVER is freely available to all researchers who sign a data usage agreement, and we hope that it will accelerate the integration of AI into healthcare settings, ultimately leading to improved patient outcomes.

## INTRODUCTION

In 2009, the Health Information Technology for Economic and Clinical Health (HITECH) Act was enacted to promote the adoption of healthcare information technology in hospitals. This act includes incentives for using electronic health record (EHR) systems[1]. The passage of the HITECH Act has resulted in widespread hospital EHR adoption with 80.5% of hospitals in the US using an EHR system, as of 2015[2]. The increased adoption of EHR systems and subsequent rise in digitally available healthcare data has resulted in a newfound ability to perform predictive modeling on healthcare data using artificial intelligence (AI), primarily in the form of machine learning. Applications of AI in a healthcare setting include providing more accurate diagnoses, recommending treatment plans, predicting patient outcomes, tracking patient engagement and adherence, reducing the burden of administrative tasks, among others [3,4,5,6,7,8, 9,10,11,12,13]. Despite many publications showing AI algorithms to be very successful in retrospective healthcare studies, there is a very limited amount of freely and publicly available medical data for researchers to work with, to develop and benchmark predictive and other methods in a reproducible manner.

To address this issue, we present and release a new repository we have constructed over the years called MOVER: Medical Informatics Operating Room Vitals and Events Repository. This repository contains data associated with hospital visits for patients undergoing surgery at the University of California, Irvine (UCI) Medical Center. The data included in MOVER was collected over seven years and contains comprehensive EHR and high fidelity physiological waveforms for patients who underwent surgery at UCI. These records include general information about each patient and their medical history, and specific information regarding the surgical procedure being performed including: medicines used, lines or drains used, and post-operative complications. The repository includes 58,799 unique patients with data from 83,468 surgeries. MOVER is freely available for download for all researchers who sign a data usage agreement and is intended to advance a wide variety of healthcare research and serve as a resource to evaluate new clinical decision support and monitoring algorithms.

## MATERIALS and METHODS

This study was approved by the Institutional Review Boards at the UCI Medical Center and on the main UCI campus (#2021-6488). Requirement for individual patient consent was waived because we removed or deidentified all protected health information (PHI) in a Health Insurance Portability and Accountability Act (HIPAA) compliant manner.

### Patient Population

This first release of MOVER includes all adult patients who underwent surgery at the UCI Medical Center from 2015–2022. The UCI Medical Center is the only level I trauma center in Orange County, California, a burn treatment center, and a National Cancer Institute-designated comprehensive cancer center. In addition, the UCI Douglas Hospital has some of the most technologically advanced surgical suites including state-of-the-art endovascular hybrid suites and intraoperative computed tomography (CT) and magnetic resonance imaging (MRI) suites.

### Data Acquisition and EHRs

The data acquisition process did not interfere with the clinical care of patients or methods of monitoring. Data for patients who underwent surgery were captured from two different sources. First, high-fidelity waveforms (EKG, pulse oximetry, and arterial line if present) from all of the operating rooms were captured in real-time using Bernoulli Health’s hardware & software platform (Bernouilli, Cardiopulmonary Corp., New Haven, CT). All of the waveforms were saved to a server on the medical center’s network organized by source location (operating room) and date/time. Subsequently, the medical center’s IT team delivered a data extract from the hospital EHR system from 2015-2022. For the years 2015-2017 the EHR system used was the Surgical Information Systems (Surgical Information Systems (SIS), Alpharetta, GASIS) and from 2017-2022 the EPIC EHR system was used (EPIC, Verona, WI). For this reason MOVER includes two datasets: the first contains two years of data from the SIS EHR system (SIS dataset) and the second contains five years of data from the EPIC EHR system (EPIC dataset).

### Repository Development and Curation

#### Data Processing

Developing MOVER involved significant data post-processing and organization. Following delivery of the data extract, the start and end time of each case was used to extract the appropriate waveform data for that case (based on location and date/time) and link it to the case. The EHR data for both datasets was then restructured and organized into logical tables (comma separated value (csv) files) for simplicity and to help facilitate data analyses. The SIS dataset has a single identifier representing each surgery, surgeries are not linked to patients and therefore it is not possible to track patients temporally across surgeries using this dataset. The raw EHR data for the EPIC dataset contained several redundant identifiers for patients and patient visits that differed between tables. To simplify this, the number of patient/event identifiers in the data were reduced to just two: the patient identifier and the patient visit identifier. This identification system allows patients to be tracked over time if they have multiple surgeries.

#### Deidentification and HIPAA Compliance

Under HIPAA Privacy Rule, all patient identifiers were removed or deidentified. For deidentification, all patient identifiers and patient visit identifiers were encoded via one-way hash functions. Additionally, PHI was removed from free-text using regular expressions and manually reviewed to ensure that all PHI was removed. Patient ages were capped at 90, so any patient with a recorded age of more than 90 years old was set to 90 years old. Ages were capped to protect patient anonymity because extreme ages are considered identifying. Finally, dates were shifted by a random number of days. The number of days by which to shift the data is linked to each patient to ensure that the data for a single patient is internally consistent. For example, if a patient had two surgeries two months apart in the raw data then the deidentified data would also reflect the surgeries as being two months apart. It is important to note, that this temporal consistency can only be observed for a single patient and not across distinct patients. For example in the deidentified data two patients who are listed as having surgery on the same day in reality did not necessarily have surgery on the same day or even in the same year.

### Repository Description

#### SIS Dataset

The SIS dataset includes 19,114 patients and is separated into nine tables: patient information, patient I/O (intake and output), patient vitals, patient observations, patient medications, patient laboratory measurements, patient procedure events, patient ventilator, and patient a-line (Supplementary Table S1). These tables contain patient demographics, information regarding the surgical procedure and anesthesia, laboratory data, and administered medications. This dataset is unique because in addition to waveforms, it contains high temporal resolution vital signs including: cardiac output, blood pressure, and stroke volume variation.

#### EPIC Dataset

The EPIC dataset is the larger of the two datasets, containing 39,685 patients, and is separated into ten tables: patient information, patient history, patient visit, patient medications, patient LDA (lines, drains, and airway devices), patient laboratory measurements, patient measurements, patient postoperative complications, patient procedure events, and patient coding (Table 1). Similar to the SIS dataset, the EPIC dataset includes patient demographics and specific information regarding the surgical procedure being performed including: medicines used, surgical events, and laboratory data. Although similar, a major difference between these two datasets is that the EPIC dataset contains outcome information including: postoperative complications, mortality, and if the patient was admitted to the ICU and the SIS dataset does not. Additionally, the EPIC dataset includes information about a patient’s medical history prior to surgery and their American Society of Anesthesiologists (ASA) physical status, while the SIS dataset does not. The final major difference is that the EPIC dataset includes billing codes.

**Table 1.** Description of the 10 tables included in the EPIC dataset

| EPIC Dataset Table | Description |
| --- | --- |
| Patient Information | patient demographic information including: age, sex, height, and weight; information regarding the surgery being performed including: the type of surgery, the start and end time of both the surgery and anesthesia, ASA status, and discharge disposition |
| Patient History | patient's health history including all patient diagnoses available in the EHR |
| Patient Visit | diagnosis and diagnosis code for a particular visit |
| Patient Medications | medications which were prescribed to patients including: the dose of the medication, the time the medication was administered or prescribed, and the medication route |
| Patient LDA | description of lines, drains, and airway devices used on the patient, the time of placement, the time of removal, and location of placement |
| Patient Labs | labs ordered for a patient, the corresponding observed measurements, and the reference measurements for each lab |
| Patient Measurements | all preoperative and postoperative patient measurements including: vitals, intake and output, pain levels, complications, and disposition |
| Patient Postoperative Complications | the type of complication and a free text note field outlining more specific details |
| Patient Procedure Events | preoperative, perioperative, and postoperative procedural events and the corresponding time of the event. |
| Patient Coding | patient billing codes |

### Repository Distribution and Documentation

MOVER is available for download at: https://mover.ics.edu, It can be downloaded by anyone who signs a data usage agreement (DUA), to restrict traffic to legitimate researchers. The website landing page has three buttons that lead to corresponding pages: documentation, data download, and article. The documentation page outlines the content of each downloadable table including: the meaning of each column, an explanation of the possible values of each column (where applicable), and the unit of measurement for each column (where applicable). On the data download page, users can sign the DUA and download the SIS and EPIC datasets. The paper page displays this publication and the citation to use when citing MOVER.

## Results and Discussion

MOVER includes 58,799 unique patients with data from 83,468 surgeries.

### SIS Dataset

The SIS dataset is the smaller of the two datasets with 19,114 patients and surgeries. Supplementary Table S2 shows summary statistics and patient demographics for all surgeries in the SIS dataset. The SIS dataset does not contain outcome information however, it does include high temporal resolution vital signs which would be invaluable for making real-time predictions to assist anesthesiologists.

### EPIC Dataset

The EPIC dataset makes up the majority of the repository, with 39,685 patients and 64,354 surgeries. Table 2 shows summary statistics and patient demographics for all surgeries in the EPIC dataset. Of the 64,354 surgeries we can see, for instance, that the average patient age was 55 and that the average length of stay was 7 days. Additionally, looking at the ASA scores we see a diverse distribution of scores with a mode of 3. The ASA score is a system used to represent a patient’s pre-anesthesia medical comorbidities, with a higher score representing a patient in worse health. Having a diverse distribution of ASA scores in this dataset shows that patients undergoing surgery are in varying degrees of health, which makes this dataset more generalizable than datasets exclusively containing patients in critical condition.

**Table 2.**
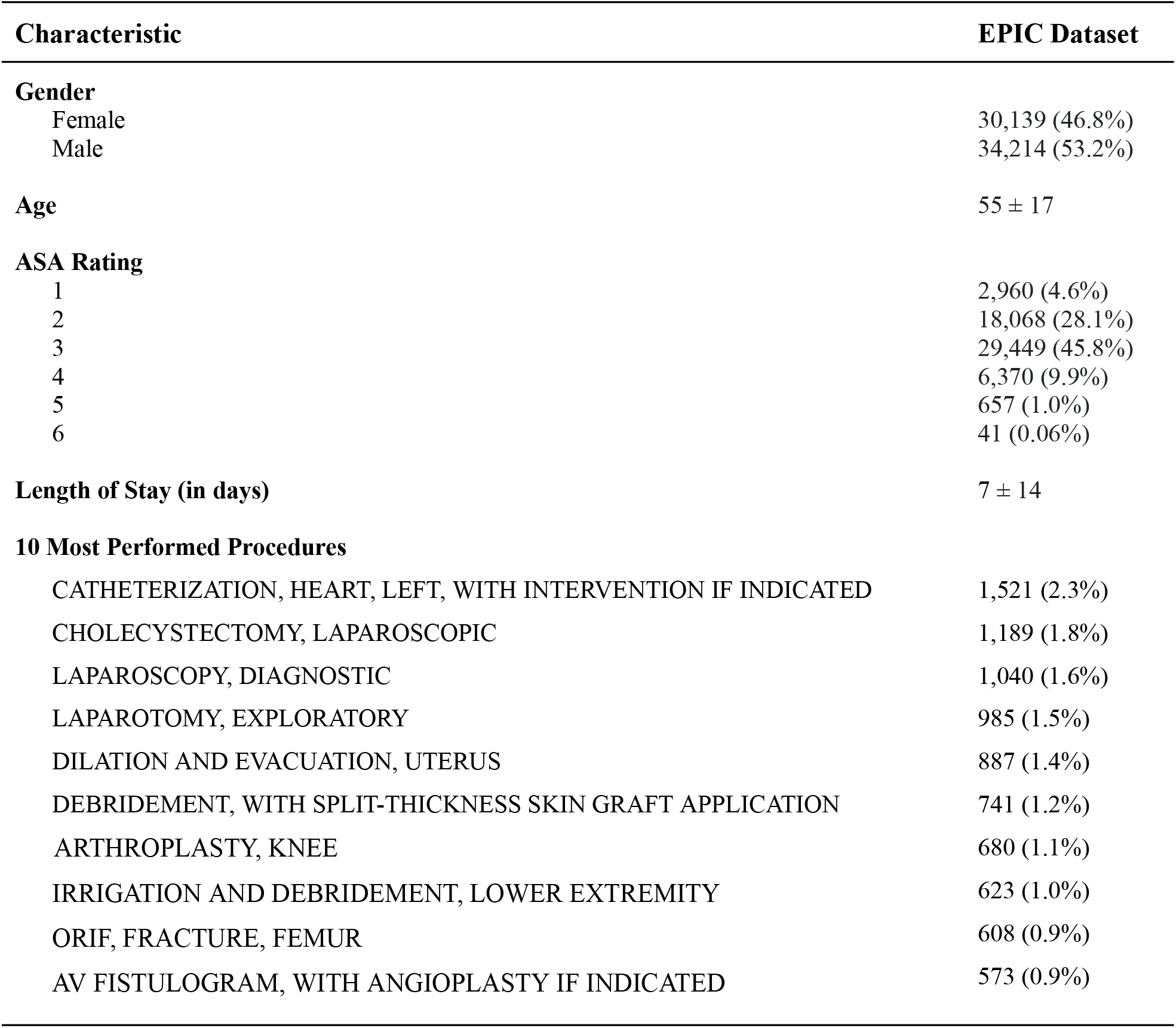
Characterization of the EPIC Dataset reported as number of records (%) or Mean ± SD

Table 3. characterizes the outcomes available in the EPIC dataset in MOVER. This characterization is useful to investigators to get an idea of what predictions they can make using MOVER. In the EPIC dataset, 45.3% of patients are transferred to the ICU after surgery and there is a 1.6% mortality rate. Table 3 also shows the percentages of the 11 classes of post-operative complications. Each postoperative complication is assigned to a class and more specific details surrounding the complication can be found in the associated free-text. Investigators would be able to use these outcomes individually for specific outcome prediction, or use them in combination to understand what factors contribute to a bad outcome of any kind.

**Table 3.** Characterization of the EPIC Dataset Outcomes reported as number of records (%)

| **Outcome** | **EPIC Dataset** |
| --- | --- |
| **Transfer to the ICU** | 29,131 (45.3%) |
| **Death** | 1,023 (1.6%) |
| **Postoperative Complications** |  |
| Other | 1,093 (1.7%) |
| Cardiovascular | 861 (1.3%) |
| Respiratory | 735 (1.1%) |
| Airway | 373 (0.6%) |
| Metabolic | 154 (0.2%) |
| Neurological | 147 (0.2%) |
| Administrative | 118 (0.2%) |
| Injury/Infection | 117 (0.2%) |
| Medication | 94 (0.1%) |
| Regional | 60 (0.1%) |
| Chronic Pain | 32 (0.05%) |

### Comparison With Other Publicly Available Clinical Repositories

To the best of our knowledge MOVER is the only freely available public data repository that contains EHR and high-fidelity physiological waveforms data for patients undergoing surgery. There are several other kinds of medical datasets that have been made publicly available including: MIMIC-IV, some of the datasets in the UCI Machine Learning Repository, and n2c2: National NLP Clinical Challenges [14,15,16,17,18]. The patient population included in MIMIC-IV are patients who were admitted to the ICU or emergency department, limiting algorithms using this data to focus on patients in critical condition. While MIMIC-IV contains more patients than MOVER, MOVER is a larger repository than MIMIC-IV containing ∼19GB of EHR data and ∼400GB of waveform data as compared to MIMIC-IV which contains ∼7GB of EHR data and ∼12GB of waveform data. The UCI Machine Learning Repository has a very limited number of small clinical datasets focusing on very specific health issues, such as diabetes, and does not include complete EHR data. The n2c2 exclusively contains unstructured text and therefore can only be used for natural language processing applications. We believe that the publication of MOVER will help address some of these limitations and complement these other publicly available datasets.

## Data Availability

All data associated with this work are available at:
https://mover.ics.uci.edu

## Acknowledgements

Work in part supported by NIH R01EB029751 (Cannesson, Baldi, and Rinehart).

## Supplementary Materials

**Table S1.** Description of the nine tables included in the SIS dataset

| SIS Dataset Table | Description |
| --- | --- |
| Patient Information | patient demographic information including: age, sex, height, and weight; information regarding the surgery being performed including: the type of surgery, the start and end time of the surgery |
| Patient Medications | medications which were prescribed to patients including: the dose of the medication, the unit of measurement for the dose, and the time the medication was administered or prescribed |
| Patient Labs | labs ordered for a patient, the corresponding observed measurements, and the time at which each measurement was taken |
| Patient I/O | patient fluid input and output information including: type of fluid, volume of fluid, and time of fluid input and or output |
| Patient Vitals | vital signs including HR from EKG, HR from pulse oximetry, non-invasive SBP, non-invasive MAP, non-invasive DBP, and SPO2 |
| Patient Observations | high temporal resolution vital signs including: CO, measurements for the femoral arterial line, measurements from the radial arterial line, SVV, cerebral oximetry, intracranial pressure mean, train of four, and several others. |
| Patient Ventilator | ventilator information including: anesthetic agent, anesthetic agent inspired fraction concentration, respiratory tidal volume, respiratory rate, airway positive inspiratory pressure, inspired fraction of nitrous oxide, and end-tidal fraction of nitrous oxide |
| Patient A-line | patient a-line placement time, location, and laterality |
| Patient Procedure Events | preoperative, perioperative, and postoperative procedural events and the corresponding time of the event |

**Table S2.** Characterization of the SIS Dataset reported as number of records (%) or Mean ± SD

| Characteristic | SIS Dataset |
| --- | --- |
| **Gender** |  |
| Female | 9,710 (50.8%) |
| Male | 9,400 (49.2%) |
| **Age** | 52 ± 19 |
| **Length of Surgery (in hours)** | 134 ± 118 |

## REFERENCES

[1] Everson, Jordan, et al. “Reconsidering Hospital EHR Adoption at the Dawn of Hitech: Implications of the Reported 9% Adoption of a ‘Basic’ EHR.” Journal of the American Medical Informatics Association, vol. 27, no. 8, 2020, pp. 1198–1205., https://doi.org/10.1093/jamia/ocaa090.

[2] Adler-Milstein, Julia, et al. “Electronic Health Record Adoption in US Hospitals: The Emergence of a Digital ‘Advanced Use’ Divide.” Journal of the American Medical Informatics Association, vol. 24, no. 6, 2017, pp. 1142–1148., https://doi.org/10.1093/jamia/ocx080.

[3] Davenport, Thomas, and Ravi Kalakota. “The Potential for Artificial Intelligence in Healthcare.” Future Healthcare Journal, vol. 6, no. 2, 2019, pp. 94–98., https://doi.org/10.7861/futurehosp.6-2-94.

[4] Lee, Christine K., et al. “Development and Validation of a Deep Neural Network Model for Prediction of Postoperative in-Hospital Mortality.” Anesthesiology, vol. 129, no. 4, 2018, pp. 649–662., https://doi.org/10.1097/aln.0000000000002186.

[5] Lilot, M., et al. “Variability in Practice and Factors Predictive of Total Crystalloid Administration during Abdominal Surgery: Retrospective Two-Centre Analysis.” British Journal of Anaesthesia, vol. 114, no. 5, 2015, pp. 767–776., https://doi.org/10.1093/bja/aeu452.

[6] Hill, Brian L., et al. “An Automated Machine Learning-Based Model Predicts Postoperative Mortality Using Readily-Extractable Preoperative Electronic Health Record Data.” British Journal of Anaesthesia, vol. 123, no. 6, 2019, pp. 877–886., https://doi.org/10.1016/j.bja.2019.07.030.

[7] Lee, Christine K., et al. “Development and Validation of an Interpretable Neural Network for Prediction of Postoperative in-Hospital Mortality.” Npj Digital Medicine, vol. 4, no. 1, 2021, https://doi.org/10.1038/s41746-020-00377-1.

[8] Cannesson, Maxime, et al. “Machine Learning of Physiological Waveforms and Electronic Health Record Data to Predict, Diagnose and Treat Haemodynamic Instability in Surgical Patients: Protocol for a Retrospective Study.” BMJ Open, vol. 9, no. 12, 2019, https://doi.org/10.1136/bmjopen-2019-031988.

[9] Kopitar, Leon, et al. “Early Detection of Type 2 Diabetes Mellitus Using Machine Learning-Based Prediction Models.” Scientific Reports, vol. 10, no. 1, 2020, https://doi.org/10.1038/s41598-020-68771-z.

[10] Datta, Arghya, et al. “Machine Learning Liver-Injuring Drug Interactions with Non-Steroidal Anti-Inflammatory Drugs (NSAIDs) from a Retrospective Electronic Health Record (EHR) Cohort.” PLOS Computational Biology, vol. 17, no. 7, 2021, https://doi.org/10.1371/journal.pcbi.1009053.

[11] Lauritsen, Simon Meyer, et al. “Explainable Artificial Intelligence Model to Predict Acute Critical Illness from Electronic Health Records.” Nature Communications, vol. 11, no. 1, 2020, https://doi.org/10.1038/s41467-020-17431-x.

[12] Baldi, P. (2021). Deep Learning in Science. Cambridge: Cambridge University Press. doi:10.1017/9781108955652

[13] Meng, Chuizheng & Trinh, Loc & Xu, Nan & Enouen, James & Liu, Yan. (2022). Interpretability and fairness evaluation of deep learning models on MIMIC-IV dataset. Scientific Reports. 12. 10.1038/s41598-022-11012-2.

[14] Johnson, A., Bulgarelli, L., Pollard, T., Horng, S., Celi, L. A., & Mark, R. (2023). MIMIC-IV (version 2.2). PhysioNet. https://doi.org/10.13026/6mm1-ek67.

[15] Johnson, A.E.W., Bulgarelli, L., Shen, L. et al. MIMIC-IV, a freely accessible electronic health record dataset. Sci Data 10, 1 (2023). https://doi.org/10.1038/s41597-022-01899-x

[16] Goldberger, A., Amaral, L., Glass, L., Hausdorff, J., Ivanov, P. C., Mark, R., … & Stanley, H. E. (2000). PhysioBank, PhysioToolkit, and PhysioNet: Components of a new research resource for complex physiologic signals. Circulation [Online]. 101 (23), pp. e215–e220.

[17] Dua, D. and Graff, C. (2019). UCI Machine Learning Repository [http://archive.ics.uci.edu/ml]. Irvine, CA: University of California, School of Information and Computer Science.

[18] “DBMI Data Portal.” DBMI Portal, https://portal.dbmi.hms.harvard.edu/.

